# Optimizing Large Language Models for Discharge Prediction: Best Practices in Leveraging Electronic Health Record Audit Logs

**DOI:** 10.1101/2024.09.12.24313594

**Authors:** Xinmeng Zhang, Chao Yan, Yuyang Yang, Zhuohang Li, Yubo Feng, Bradley A. Malin, You Chen

## Abstract

Electronic Health Record (EHR) audit log data are increasingly utilized for clinical tasks, from workflow modeling to predictive analyses of discharge events, adverse kidney outcomes, and hospital readmissions. These data encapsulate user-EHR interactions, reflecting both healthcare professionals’ behavior and patients’ health statuses. To harness this temporal information effectively, this study explores the application of Large Language Models (LLMs) in leveraging audit log data for clinical prediction tasks, specifically focusing on discharge predictions. Utilizing a year’s worth of EHR data from Vanderbilt University Medical Center, we fine-tuned LLMs with randomly selected 10,000 training examples. Our findings reveal that LLaMA-2 70B, with an AUROC of 0.80 [0.77-0.82], outperforms both GPT-4 128K in a zero-shot, with an AUROC of 0.68 [0.65-0.71], and DeBERTa, with an AUROC of 0.78 [0.75-0.82]. Among various serialization methods, the first-occurrence approach—wherein only the initial appearance of each event in a sequence is retained—shows superior performance. Furthermore, for the fine-tuned LLaMA-2 70B, logit outputs yield a higher AUROC of 0.80 [0.77-0.82] compared to text outputs, with an AUROC of 0.69 [0.67-0.72]. This study underscores the potential of fine-tuned LLMs, particularly when combined with strategic sequence serialization, in advancing clinical prediction tasks.

## Introduction

The audit log data of electronic health record (EHR) systems passively document metadata that objectively captures system users’ granular interactions with the EHR interface. The audit log data contains various information, including, but not limited to, user ID, action performed (characterized by free-text descriptions), patient ID of the accessed EHR, and time of access. This data, mandated by the Health Insurance Portability and Accountability Act of 1996^1^, was originally designed for forensic purposes and to enhance patient privacy protections. However, it has since garnered substantial interest in its potential to serve as a resource for modeling and measuring clinical workflow and burden. EHR audit logs have been successfully used for measuring clinician engagement with EHRs^2^, characterizing clinical workflow patterns^3,4^, examining physicians’ workload to identify potential burnout^5^, and analyzing disparities in care delivery^6^.

Notably, due to their ability to capture a healthcare professional’s real-time assessment of a patient’s health status, EHR audit logs have proven effective in improving clinical outcome prediction tasks^7,8^. For instance, in a recent study on predicting inpatient discharge within the next 24 hours, researchers found that incorporating healthcare professionals’ interactions with the EHR system enhanced prediction performance, increasing the area under the receiver operating characteristic curve (AUC) from 0.86 to 0.92^7^. In another study, integrating EHR audit logs into a machine learning model for forecasting major adverse kidney events within 120 days after ICU admission in acute kidney injury patients significantly improved AUC from 0.79 to 0.88^8^. Additionally, for predicting all-cause hospital readmission within 30 days after discharge in acute stroke patients, the AUC increased from 0.63 to 0.74.

To date, most relevant studies have focused on incorporating EHR audit log data as structured features within traditional machine learning models. User actions described in the text were usually categorized and represented as atemporal features that are encoded in a vectorized format, which does not capture the semantic meaning of each action. However, the rich temporal and semantic information contained in EHR audit logs provides an opportunity to further inform clinical prediction tasks. Yet, harnessing the temporal aspects of audit log sequences is a non-trivial endeavor. In a prior study, researchers found that a bidirectional RETAIN-based model—a variation of a recurrent neural network (RNN) with a custom embedding generated by Word2Vec to capture the temporal relationship of EHR audit logs—performed worse than standard tree-based models (specifically, Light Gradient-Boosting Machine (LGBM))^7^. In the Word2Vec approach, instead of using the sequence of textual descriptions, each audit log action type was encoded as a unique ID. Therefore, one of the likely reasons for this underperformance is that the autoregressive model, along with the word embedding strategy, was unable to adequately represent the patterns in the sequence of textual descriptions.

This discrepancy in performance highlights the limitations of traditional machine learning models and word embedding techniques for handling complex sequence modeling tasks. Recently, large language models (LLMs) have led to breakthroughs in many areas of natural language processing^9,10^. To the best of our knowledge, there is currently no research on the utilization of LLMs for analyzing audit log data. Thus, we conducted an empirical study to evaluate an LLM’s ability to process EHR audit log user action sequences and their performance in real-world clinical prediction tasks using these sequences. Specifically, we aimed to answer the following questions. First, how effectively can LLMs encode EHR audit log user action sequences and leverage the extracted information for predicting patient outcomes? Second, how does the performance of using an LLM for text-level classification, where the model generates textual outputs of discharge status, compare to logit-level classification, where the model is fine-tuned to predict discharge status by mapping its hidden states to binary classes? Third, how do different methods for converting structured EHR audit log user action sequences into natural language inputs (i.e., serialization) affect performance? As a case study, we evaluate GPT-4 128k and LLaMA-2 on the prediction of 24-hour inpatient discharge.

## Related Work

The transformer-based architecture of LLMs has shown exceptional capability in complex nature language processing tasks, particularly in handling long-range dependencies^11^. Pre-trained LLMs, such as GPT-3 (175B parameters), GPT-4 128k (allegedly 1.76T parameters), and LLaMA-2 (7B, 13B, and 70B parameters), have demonstrated impressive performance across various tasks—even in zero-shot settings (i.e., without any task-specific training data)—such as question answering^12^ and information retrieval^13^. However, studies have shown that, for tasks with data types that LLMs have not been explicitly pre-trained on (e.g., tabular data), zero-shot performance can be worse than traditional machine learning models, such as Logistic Regression^14^. An LLM’s ability to perform clinical outcome prediction tasks is often limited due to the lack of real-world healthcare data in its training dataset, as well as a lack of alignment with clinical outcome prediction goals^15^. Few-shot in-context learning has demonstrated effectiveness in aligning LLMs with clinical prediction tasks, such as drug pair synergy predictions in rare tissues, where data is limited^10^. Furthermore, when computational resources and appropriate data are made available, supervised LLM fine-tuning may further improve performance for specific clinical tasks^9^.

## Methods

This retrospective study was approved by the institutional review board (IRB) at Vanderbilt University Medical Center (VUMC) under IRB#191892.

### Data

This study utilized EHRs and EHR access log data from VUMC, a large academic medical center in Nashville, Tennessee. VUMC migrated to the Epic EHR system in 2018. We collected data from all adult inpatient visits in 2019, focusing on those with complete demographic information, a length of stay greater than 24 hours, and discharges to either home or care facilities. For patients with more than one inpatient stay in 2019, we retained the first visit to mitigate potential dominating effects from this subgroup. We focused on the prediction task described by *Zhang et al*.^7^, which aims to forecast whether a patient would be discharged within the next 24 hours, with predictions made daily at 2:00 pm. We divided each inpatient visit into daily instances, each containing EHR audit log information from the preceding 24 hours. This yielded data for 31,688 unique patients and 152,490 instances.

### EHR audit log data

EHR audit log data are an extensive and detailed data resource that captures the granular interactions between healthcare professionals and the EHR system. They record every action performed by the users of the EHR system, including healthcare professionals, administrative personnel, and other authorized individuals. More specifically, EHR audit logs document who performed what action(s), in which patient’s EHR, and at what time. EHR vendors typically provide an expansive list of user actions, along with free-text descriptions, ensuring that any action taken is accurately logged with context. Table 1 provides an example of an EHR audit log user action sequence for a single patient. This example includes user ID, patient ID, user action type, and the timestamp associated with each action.

**Table 1.** An example of an EHR audit log user action sequence of a single patient.

| User ID | Patient ID | EHR audit log user action | Timestamp |
| --- | --- | --- | --- |
| U001 | 1 | Flowsheet data saved. | 2/1/2019 12:05 |
| U001 | 1 | Patient report accessed. | 2/1/2019 12:15 |
| U002 | 1 | Lab results viewed. | 2/1/2019 14:10 |
| U001 | 1 | Print out activity. | 2/2/2019 6:20 |
| U003 | 1 | Medication tab selected. | 2/2/2019 6:25 |

### Serialization methods

Considering the sensitivity of LLMs to input formats and their limitations regarding input length, this study focuses on various strategies for serializing user action sequences. We investigated three serialization methods, each varying in input length and complexity. Our analysis includes an example of a raw user action sequence, represented as A→B→B→C→B, as shown in Figure 1a.

**Figure 1.**
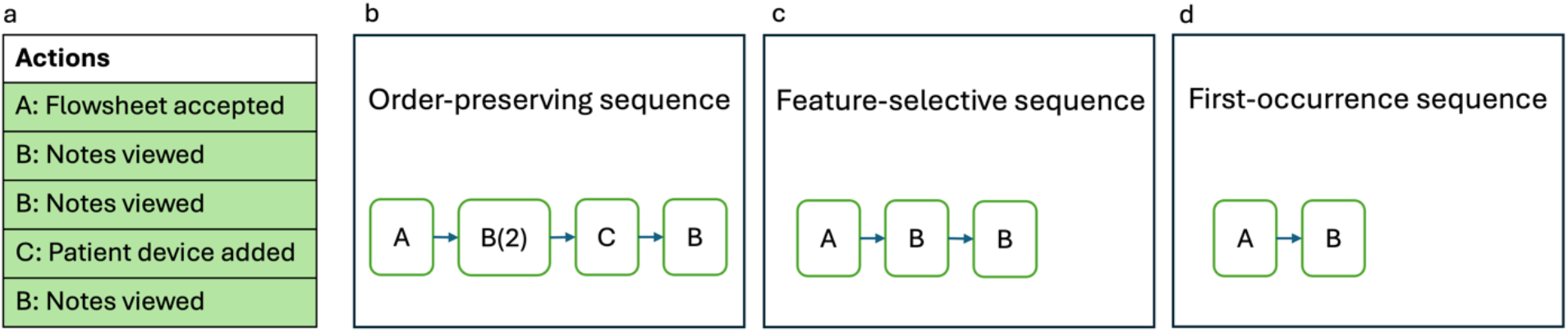
Overview of the serialization methods. **a**, An example of raw EHR audit log user action sequence for a patient within the past 24 hours. In this example, ‘Flowsheet accepted’ and ‘Notes viewed event’ actions were among the *k* most important features for discharge prediction. **b**, The order-preserving sequence serialization. **c**, The feature-selective sequence serialization. **d**, The first-occurrence sequence serialization.

1) **Order-preserving sequence serialization:** This method simplifies user action sequences while preserving their original order by removing consecutive duplicates. Each set of duplicates is replaced with a single action notation along with a count of the occurrences, as illustrated in Figure 1b.
2) **Feature-selective sequence serialization:** Building upon the order-preserving sequence approach, this method creates sequences composed of actions identified as the top *k* important features for discharge prediction as determined by LGBM in *Zhang et al*.^7^. For example, if actions A and B rank among the top *k* action types, then the sequence becomes A→B→B (Figure 1c). We considered *k* = 10 and 30 in this study.
3) **First-occurrence sequence serialization:** Using the sequence produced by the feature-selective sequence approach, this approach retains only the first occurrence of each action. For instance, the sequence is reduced to A→B (Figure 1d).

### Large Language Models for Classification

We explored two distinct paradigms for applying LLMs to discharge status prediction: 1) text-level classification and 2) logit-level classification. In the text-level classification, as shown in Figure 2a, both the input and output of an LLM are in free-text format. The output layer of the LLM maps its hidden state to a sequence of words with the highest conditional probability. We designed LLM instructions to prompt the model to produce JSON-formatted responses of either “Discharged” or “Not discharged”. These textual outputs were subsequently mapped to positive or negative predictions accordingly. This method enables the direct application of pretrained LLMs for zero-shot tasks and few-shot in-context learning tasks. In contrast, logit-level classification (Figure 2b) involves fine-tuning an LLM for a specific task to generate a numerical probability. In this paradigm, the output layer of the LLM maps hidden states to two classes and outputs the corresponding logits.

Patient demographic and clinical data were serialized using a text template format: “The column name: value”. They include 1) age, 2) gender, 3) insurance type, 4) hospital stay duration, 5) prediction day of the week, 6) admission diagnoses, and 7) historical diagnoses over the past 10 years.

**Figure 2.**
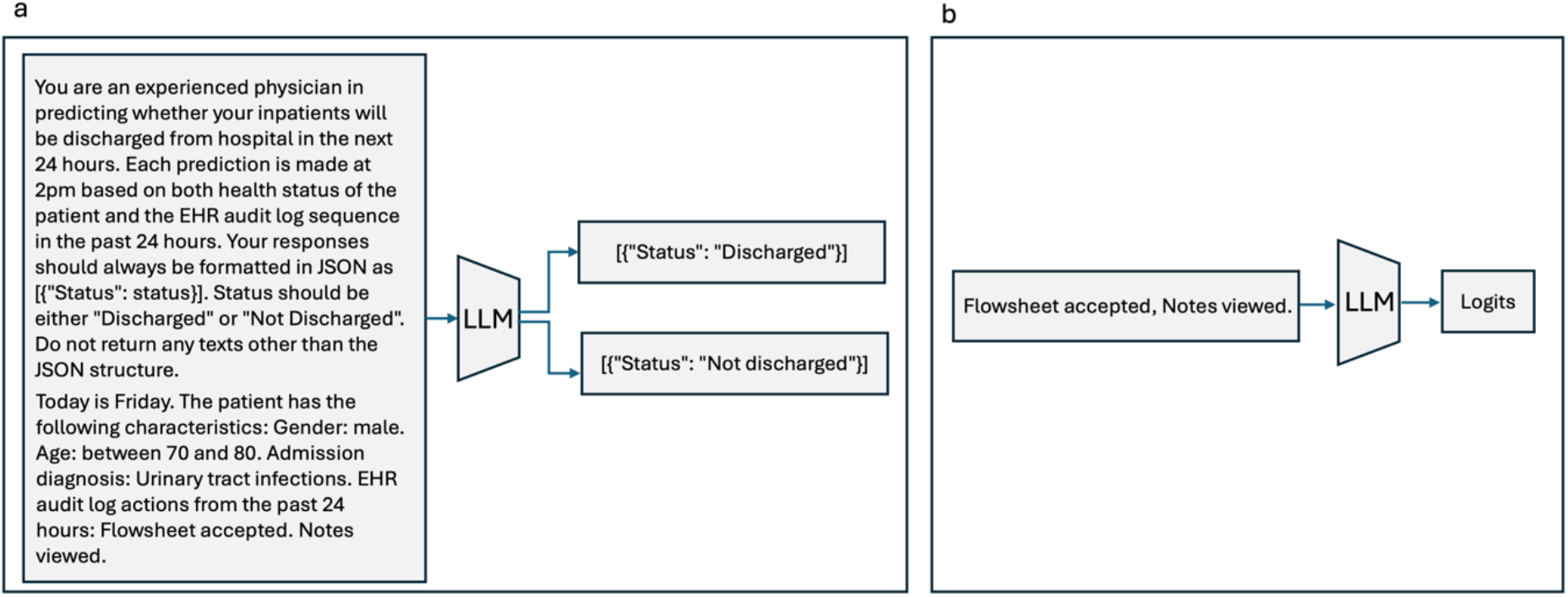
An illustration of LLM prediction paradigms. **a**, In the text-level classification task, the LLM generates text following a predefined format, using a user action sequence as the input. **b**, In the logit-level classification task, the LLM produces a set of logits as its output.

### Experiment Design

For LLM evaluation and fine-tuning, we randomly selected 10,000 instances as the training dataset and 1,500 instances as the testing dataset. We ensured no patient data appeared in both datasets. For feature selection, we utilized 85% of the instances to train an LGBM model and utilized SHapley Additive exPlanations) (SHAP) values on 10% of the instances to identify the *k* most important features. Further details on training the LGBM models and identifying important features can be found in *Zhang et al*.^7^.

To answer our research questions, we prompted GPT-4 128k with zero-shot and one-shot strategies and fine-tuned LLaMA-2 in our experiments. To evaluate LLM’s capability in understanding EHR audit log data, we conducted both zero-shot and one-shot (i.e., using one randomly selected example with a positive label from the training dataset) experiments with GPT-4 128k for the text-level classification. The prompt for GPT-4 128k consists of a user action sequence using the order-preserving sequence serialization and serialized demographic and clinical information, providing the most comprehensive information compared to non-GPT4 models.

To evaluate the performance of using an LLM for text-level classification compared to logit-level classification, we used the first-occurrence serialization with *k* = 10 in LLaMA-2 (7B and 70B models). We applied the Quantized Low Rank Adapters (QLoRA) technique, which uses 4-bit quantized models and incorporates a small set of parameters into the model for efficient fine-tuning^16^. We fine-tuned all models with a LoRA rank of 8 and alpha of 16, loading models in 4-bit using nested quantization.

To evaluate how different serialization methods affect prediction performance, we focused on the logit-level classification task and experimented with LLaMA-2 7B using both feature-selection serialization and first-occurrence serialization with different feature numbers (*k* = 10 and 30). The most effective serialization method was then applied to the LLaMA-2 70B model to examine how model sizes influence prediction performance. We did not include serialized demographic and clinical information in experiments using LLaMA-2 due to the token limits and observed performance degradation with increased context length. Additionally, to evaluate the influence of serialization methods in the zero-shot setting, we ran experiments with GPT-4 128k using only the audit log user action sequences with the first-occurrence sequence to compare the performance with GPT-4 128k model using the order-preserving sequence serialization combined with demographic and clinical information.

In addition, we conducted multiple ablation studies. First, we compared the classification performances of LLaMA-2 7B with a smaller alternative–Decoding-enhanced BERT with Disentangled Attention (DeBERTa-v3-small)^17^. Second, to assess the impact of feature selection on prediction performance, we conducted an experiment by randomly selecting 10 user action types from those that were not included in the top 50 most significant user action types. Additionally, we evaluated the performance of LGBM using all EHR audit log user actions.

We utilized two NVIDIA A100 (80GB) GPUs for our experiments. Area under the receiver operating characteristics curve (AUC) and F1 scores were used for evaluation purposes. For calculating AUC in the text-level classification setting, we converted the textual outputs from GPT-4 128k into binary numeric values (1 for “Discharged” and 0 for “Not discharged”). To calculate F1 scores in the logit-level classification setting, we selected the threshold at which the recall was approximately 0.8. We then bootstrapped the testing dataset 1,000 times with replacements to calculate the 95% confidence interval (CI) for the AUC and F1 score. All experiments with GPT-4 128k were conducted using the secured Azure OpenAI API, managed by VUMC.

## Results

Table 2 presents the summary statistics about the dataset used in this study. The average number of EHR audit log user actions was 2,231 with a standard derivation (SD) of 1,178 in the training dataset and 2,128 (SD = 1,279) in the testing dataset. The ratio of positive to negative instances was approximately 1:4 in both datasets.

**Table 2.**
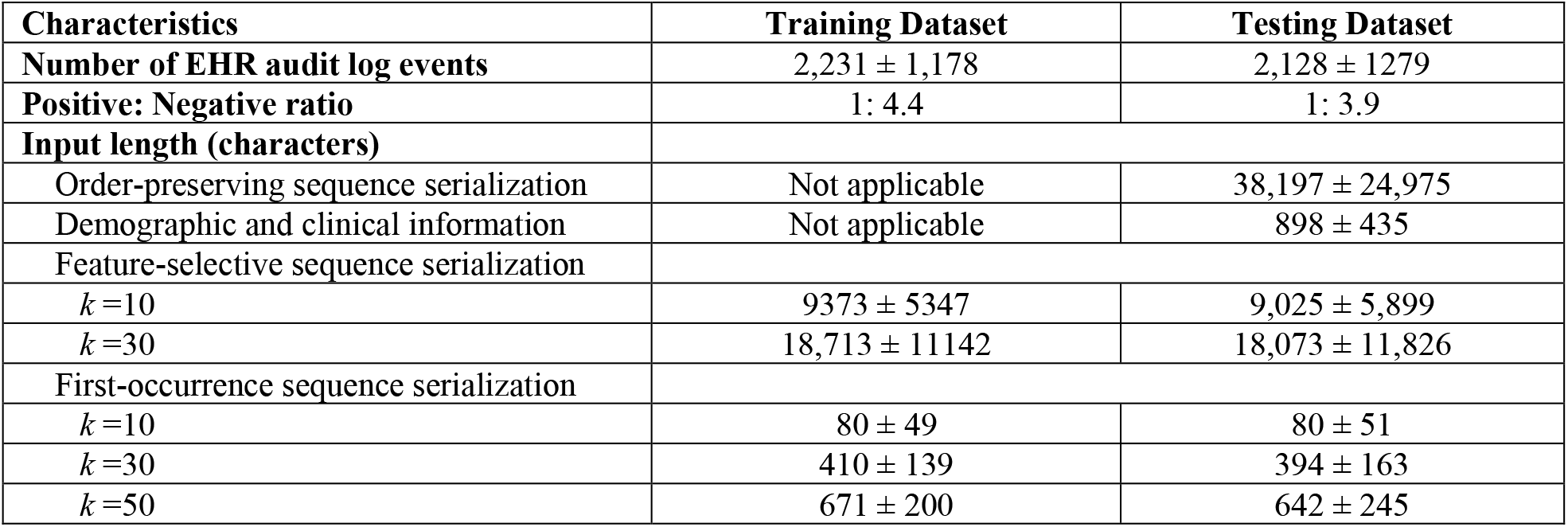
Summary of the dataset used in this study. *x* ± *y* indicates the mean ± one standard deviation.

User action sequence input lengths were measured in characters because variations exist in token counts across different models’ tokenizers. The order-preserving sequence serialization method generated the longest input with a mean (SD) input length of 38,197 (24,975) characters in the test dataset. The feature-selective sequence serialization method considerably shortened the length lengths, with a mean (SD) length of 9,025 (5,899) for *k* = 10 and 18,073 (11,826) for *k* = 30 in the test dataset. We excluded the *k* = 50 condition from the experiment because the input length at *k* = 30 has already reached the token limit of 4,096 for the LLaMA-2 models. The first-occurrence sequence serialization resulted in the most concise input sequences. The mean (SD) of the input length in the test dataset was 80 (51), 394 (163), and 642 (245) for *k* equal to 10, 30, and 50, respectively.

### LLMs’ prior knowledge of EHR audit logs

Combining the EHR audit log user action sequence of a patient and their demographic and clinical information, GPT-4 128k achieved an AUC of 0.65 and an F1 score of 0.42 in the zero-shot setting (Table 3). This suggests that GPT-4’s pretrained knowledge may not sufficiently cover EHR audit log data, as well as the predictive signals from the data for discharge prediction. When shifting to the one-shot setting, providing GPT-4 128k with a randomly selected example, the AUC reduced to 0.62 and the F1 score to 0.40, suggesting that the additional but limited context has the potential to degrade the model’s performance, particularly with the data that the models were not trained on in the pretraining stage.

**Table 3.** The performance of GPT-4 128k with zero- and one-shot settings.

| Serialization Approaches | Learning paradigm | AUC [95% CI] | F1 score [95% CI] |
| --- | --- | --- | --- |
| Order-preserving sequence + demographic and clinical information | Zero-shot | 0.65<br>[0.62-0.67] | 0.42<br>[0.39-0.46] |
|  | One-shot | 0.62<br>[0.60-0.65] | 0.40<br>[0.37-0.44] |

### Comparison between text-level and logit-level classifications

For the text-level classification task, LLaMA-2 7B, which utilized the first-occurrence sequence serialization with *k*=10, achieved an AUC of 0.62 and an F1 score of 0.40 (Table 4). LLaMA-2 70B exhibited an improved performance with an AUC of 0.69 and an F1 score of 0.54. This improvement suggests the potential benefits of larger model sizes for enhancing predictive performance after fine-tuning. However, LLaMA-2’s performance with the EHR audit log user action sequences for the textual output prediction task remained poor.

**Table 4.**
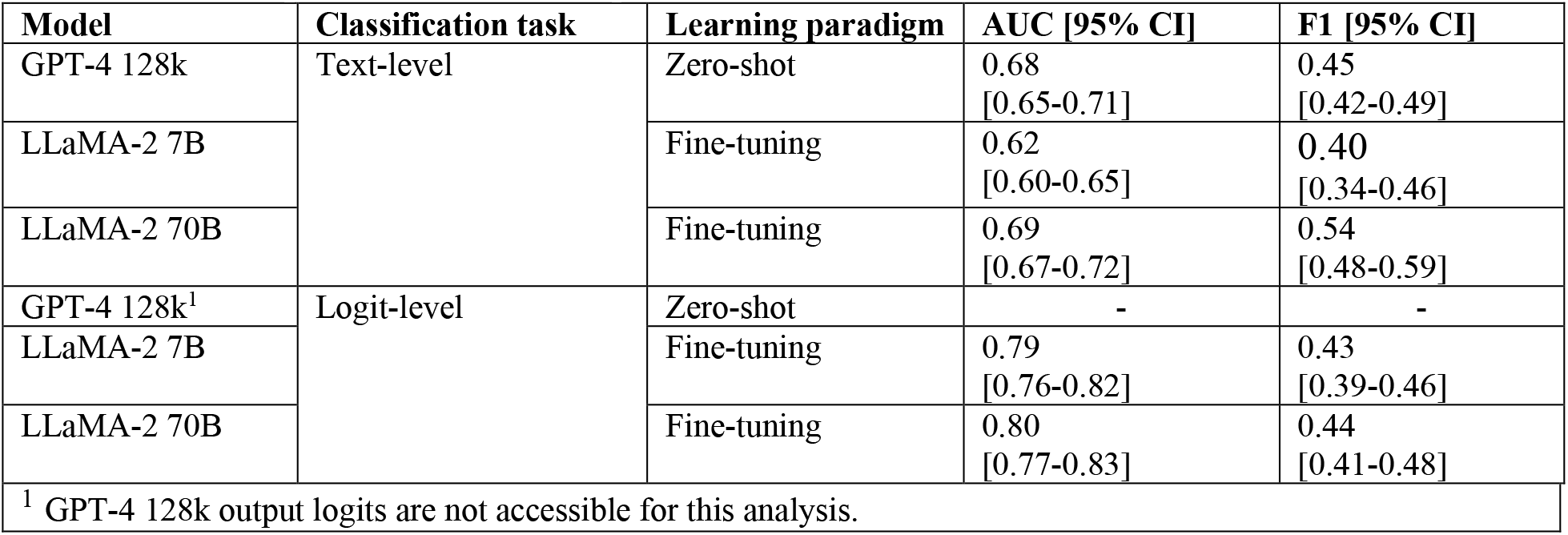
The performance comparison of GPT4-128k and LLaMA-2 in the text-level classification and logit-level classification utilizing the first-occurrence sequence serialization with *k*=10.

| Model | Classification task | Learning paradigm | AUC [95% CI] | F1 [95% CI] |
| --- | --- | --- | --- | --- |
| GPT-4 128k | Text-level | Zero-shot | 0.68<br>[0.65-0.71] | 0.45<br>[0.42-0.49] |
| LLaMA-2 7B |  | Fine-tuning | 0.62<br>[0.60-0.65] | 0.40<br>[0.34-0.46] |
| LLaMA-2 70B |  | Fine-tuning | 0.69<br>[0.67-0.72] | 0.54<br>[0.48-0.59] |
| GPT-4 128k<sup>1</sup> | Logit-level | Zero-shot | - | - |
| LLaMA-2 7B |  | Fine-tuning | 0.79<br>[0.76-0.82] | 0.43<br>[0.39-0.46] |
| LLaMA-2 70B |  | Fine-tuning | 0.80<br>[0.77-0.83] | 0.44<br>[0.41-0.48] |

<sup>1</sup> GPT-4 128k output logits are not accessible for this analysis.

Additionally, we tested GPT-4 128k using the same serialization method (i.e., first-occurrence sequence serialization with *k* = 10) in a zero-shot setting. In this situation, GPT-4 128k achieved an AUC of 0.68 and an F1 score of 0.45, which surpassed its performance with a more comprehensive, though lengthier, input (i.e., order-preserving sequence + demographic and clinical information), as shown in Table 3. This observation indicates that while GPT4-128k enables a larger context window, its predictive capability degrades as the length of the input context increases, highlighting a phenomenon known as the “lost in the middle” problem^18^.

When utilizing LLaMA-2 for the logit-level classification, we observed a substantial performance improvement compared to the text-level classification. With the same serialization method (i.e., first-occurrence sequence serialization with *k*=10), LLaMA-2 7B and 70B models achieved an AUC of 0.79 and 0.80 and F1 scores of 0.43 and 0.44, respectively, which contrast with an AUC of 0.68 achieved by the zero-shot GPT-4 128k model for the text-level classification task. Additionally, in contrast to the large performance gap between LLaMA-2 7B and 70B in the text-level classification task, for the logit-level classification task, 7B and 70B models performed similarly with an AUC of 0.79 and 0.80 and an F1 score of 0.43 and 0.45, respectively.

### Optimal strategies for serializing EHR audit log sequences

At the logit-level classification setting, we evaluated the performance of LLaMA-2 7B across different serialization methods for EHR audit log sequences. As shown in Table 5, in the experiments with the feature-selective sequence serialization, the results indicated that the LLM processed shorter sequences more effectively. Specifically, when using *k*=10, fine-tuned LLaMA-2 7B achieved a higher AUC of 0.69 and an F1 score of 0.41 compared to *k* = 30 (Table 5). When the sequences were further reduced with the first-occurrence sequence serialization method, fine-tuned LLaMA-2 7B exhibited comparable prediction performances across different values of *k*. Comparing first-occurrence sequence serialization and feature-selective sequence serialization with the same value of *k*, we observed that feature-selective sequence serialization, which produces more comprehensive, but lengthier, input does not yield better predictive performance.

**Table 5.** A comparison of LLaMA-2 models for logit-level classification with different serialization strategies with LGBM as a baseline.

| Model | Learning paradigm | Serialization Approaches | k | AUC [95% CI] | F1 [95% CI] |
| --- | --- | --- | --- | --- | --- |
| LGBM | Training | Not applicable | - | 0.86<br>[0.84-0.89] | 0.53<br>[0.49-0.57] |
| LLaMA-2 7B | Fine-tuning | First-occurrence sequence serialization | 10 | 0.79<br>[0.76-0.82] | 0.43<br>[0.39-0.46] |
|  |  |  | 30 | 0.81<br>[0.77-0.84] | 0.45<br>[0.41-0.48] |
| LLaMA-2 7B | Fine-tuning | Feature-selective sequence serialization | 10 | 0.69<br>[0.66-0.72] | 0.41<br>[0.38-0.45] |
|  |  |  | 30 | 0.53<br>[0.50-0.56] | 0.35<br>[0.32-0.38] |

**Table 6.**
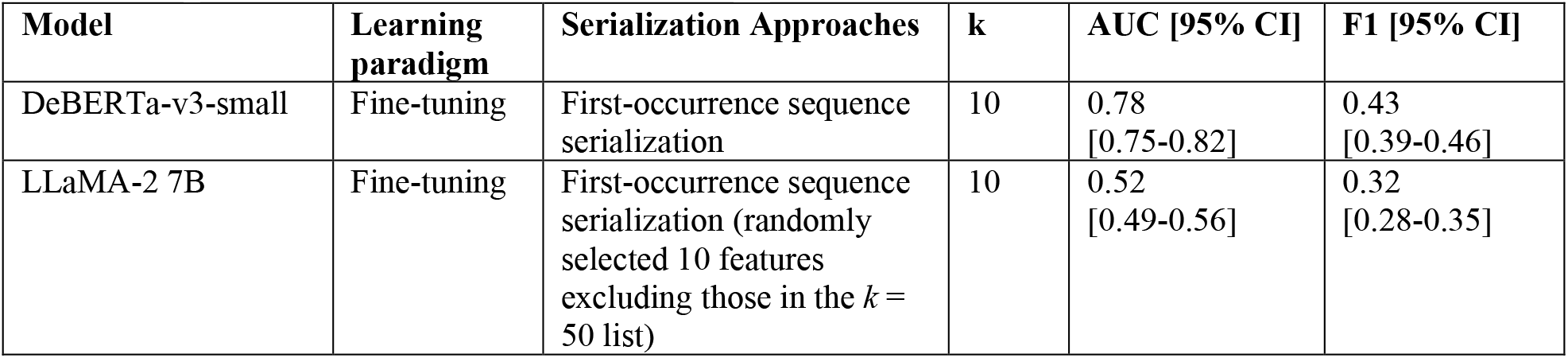
The performance of ablation studies on the logit-level classification task.

| Model | Learning paradigm | Serialization Approaches | k | AUC [95% CI] | F1 [95% CI] |
| --- | --- | --- | --- | --- | --- |
| DeBERTa-v3-small | Fine-tuning | First-occurrence sequence serialization | 10 | 0.78<br>[0.75-0.82] | 0.43<br>[0.39-0.46] |
| LLaMA-2 7B | Fine-tuning | First-occurrence sequence serialization (randomly selected 10 features excluding those in the $k = 50$ list) | 10 | 0.52<br>[0.49-0.56] | 0.32<br>[0.28-0.35] |

### Ablation studies

In the comparison of the DeBERTa model using the first-occurrence sequence serialization of 10 features with LLaMA-2 7B, the fine-tuned DeBERTa model achieved an AUC of 0.78 and an F1 score of 0.43. The performance was slightly lower than the fine-tuned LLaMA-2 7B (AUC = 0.79). This finding illustrates the robust capacity of DeBERTa for the classification task with a brief input sequence. In terms of feature selection, the fine-tuned LLaMA-2 7B model exhibited a substantial reduction in performance, as highlighted by an AUC of 0.52 and an F1 score of 0.32, indicating the critical role of feature selection strategy for reducing input sequence length.

## Discussion

Our study investigates the opportunities for adapting LLMs that are pre-trained on general domain data to medical classification tasks. In our analysis of EHR audit log user action sequences for discharge prediction, we found that GPT-4 128k, in a zero-shot setting, demonstrated poor performance. This finding is consistent with previous studies that reported weak zero-shot or very-few-shot performances by LLMs such as GPT-4 128k on medical domain tasks^14^, potentially due to their limited exposure to medical data during pretraining. This problem becomes even more prominent in our setting, which may be attributed to the absence of publicly available EHR audit logs. Our findings underscore the need for the development of medical domain-specific and data type-aware LLMs to achieve sufficiently high performance on clinical outcome prediction tasks.

Fine-tuning is a strategy to improve LLM performance by adapting them to a specific task. However, when using LLMs for classification, there exists a trade-off between leveraging the models’ prior knowledge and optimizing task-specific effectiveness. When applying LLMs for text-level classification, direct application of LLMs without fine-tuning is possible but may suffer from poor zero-shot capabilities and limited gains from one-shot in-context learning. On the other hand, adapting LLMs to function as logit-level classifiers requires model fine-tuning on a specific task and thus can achieve better performance than pure inference at the expense of computational costs. Thus, it is in the best interests of practitioners to assess both approaches based on their specific datasets and classification tasks.

Moreover, our experiments demonstrated that the serialization strategy has a non-trivial impact on an LLM’s performance. As shown in our results, serialization methods greatly influence an LLM’s performances in both zero-shot and fine-tuning settings, where shorter inputs generally result in higher prediction performance. In the zero-shot setting, GPT-4 128k performed better with shorter inputs as opposed to more comprehensive but lengthier inputs. In fine-tuning experiments, we observed that LLaMA-2 models using first-occurrence serialization inputs outperformed those using feature-selection serialization inputs. These results suggested that effectively summarizing EHR audit log user action sequences into concise formats that capture essential information can substantially improve the predictive capabilities of LLMs.

In the comparison of LLMs with traditional tree-based models, we observed that the tree-based boosting model outperformed the fine-tuned LLMs. When comparing LLMs to a small-size pre-train language model, the fine-tuned DeBERTa achieved a slightly lower performance than the fine-tuned LLaMA-2 model but much higher performance than GPT-4 128k in the zero-shot setting. These results align with prior observations in the literature where LGBM and XGBoost outperformed LLMs on various classification tasks^14^. The inefficacy of general domain LLMs highlights the importance of pretraining on medical domain data for improvement in medical classification tasks.

Recently, LLMs specific to the medical domain have been developed, such as Med-PaLM 2^19^ and HuatuoGPT^20^, which are trained or fine-tuned with medical-domain knowledge to better align with clinical tasks. These models have shown great promise in medical question-answering tasks, including US Medical Licensing Examination-style questions. Some clinical-focused LLMs^21-23^ were trained on EHRs, such as MIMIC-III^24^, to accomplish clinical prediction tasks. Despite this progress, further investigation is necessary to compare the performance of medical-domain LLMs in classification tasks with traditional models. Furthermore, the broader impact and usefulness of medical-domain LLMs to the health systems remain unevaluated^25^.

## Limitations and Conclusion

There are several limitations of this study that we wish to acknowledge. First, we did not evaluate the performance of fine-tuned OpenAI models. Future studies should explore how the performance of OpenAI models is influenced by the number of fine-tuning examples. Second, our study focused only on a single task of discharge prediction. To more comprehensively evaluate the potential of LLMs with EHR audit logs, additional clinical prediction tasks should be evaluated. Third, future studies should explore variants of LLaMA with extended context length, such as LongLLaMA^26^ to determine their effectiveness in handling longer sequences. Finally, the implementation of our proposed method requires access to GPU resources, which are increasingly accessible through cloud computing services.

## Data Availability

All data produced in the present study are available upon reasonable request to the authors.

## Acknowledgments

This research was supported, in part, by the National Center for Advancing Translational Sciences of the National Institutes of Health under Award Number UL1TR002243. The content is solely the responsibility of the authors and does not necessarily represent the official views of the National Institutes of Health.

## Notes

### Competing Interest Statement

The authors have declared no competing interest.

## References

1. National Research Council (US) Committee on Maintaining Privacy and Security in Health Care Applications of the National Information Infrastructure. For the Record Protecting Electronic Health Information [Internet]. Washington (DC): National Academies Press (US); 1997 [cited 2024 Mar 18]. Available from: http://www.ncbi.nlm.nih.gov/books/NBK233429/

2. Adler-Milstein J, Adelman JS, Tai-Seale M, Patel VL, Dymek C. EHR audit logs: A new goldmine for health services research? J Biomed Inform. 2020 Jan;101:103343.

3. Chen B, Alrifai W, Gao C, Jones B, Novak L, Lorenzi N, et al. Mining tasks and task characteristics from electronic health record audit logs with unsupervised machine learning. J Am Med Inform Assoc. 2021 Jun 12;28(6):1168–77.

4. Zhang X, Zhao Y, Yan C, Derr T, Chen Y. Inferring EHR Utilization Workflows through Audit Logs. AMIA Annu Symp Proc. 2023 Apr 29;2022:1247–56.

5. Kannampallil T, Adler-Milstein J. Using electronic health record audit log data for research: insights from early efforts. J Am Med Inform Assoc. 2023 Jan 1;30(1):167–71.

6. Yan C, Zhang X, Yang Y, Kang K, Were MC, Embí P, et al. Differences in Health Professionals’ Engagement With Electronic Health Records Based on Inpatient Race and Ethnicity. JAMA Netw Open. 2023 Oct 2;6(10):e2336383.

7. Zhang X, Yan C, Malin BA, Patel MB, Chen Y. Predicting next-day discharge via electronic health record access logs. J Am Med Inform Assoc JAMIA. 2021 Nov 25;28(12):2670–80.

8. Bhaskhar N, Ip W, Chen JH, Rubin DL. Clinical outcome prediction using observational supervision with electronic health records and audit logs. J Biomed Inform. 2023 Nov 1;147:104522.

9. Guevara M, Chen S, Thomas S, Chaunzwa TL, Franco I, Kann BH, et al. Large language models to identify social determinants of health in electronic health records. Npj Digit Med. 2024 Jan 11;7(1):1–14.

10. Li T, Shetty S, Kamath A, Jaiswal A, Jiang X, Ding Y, et al. CancerGPT for few shot drug pair synergy prediction using large pretrained language models. Npj Digit Med. 2024 Feb 19;7(1):1–10.

11. Vaswani A, Shazeer N, Parmar N, Uszkoreit J, Jones L, Gomez AN, et al. Attention is All you Need. In: Advances in Neural Information Processing Systems [Internet]. Curran Associates, Inc.; 2017 [cited 2024 Mar 18]. Available from: https://proceedings.neurips.cc/paper_files/paper/2017/hash/3f5ee243547dee91fbd053c1c4a845aa-Abstract.html

12. Brown TB, Mann B, Ryder N, Subbiah M, Kaplan J, Dhariwal P, et al. Language Models are Few-Shot Learners [Internet]. arXiv; 2020 [cited 2024 Mar 18]. Available from: http://arxiv.org/abs/2005.14165

13. Touvron H, Martin L, Stone K, Albert P, Almahairi A, Babaei Y, et al. Llama 2: Open Foundation and Fine-Tuned Chat Models [Internet]. arXiv; 2023 [cited 2024 Mar 18]. Available from: http://arxiv.org/abs/2307.09288

14. Hegselmann S, Buendia A, Lang H, Agrawal M, Jiang X, Sontag D. TabLLM: Few-shot Classification of Tabular Data with Large Language Models. In: Proceedings of The 26th International Conference on Artificial Intelligence and Statistics [Internet]. PMLR; 2023 [cited 2023 Aug 30]. p. 5549–81. Available from: https://proceedings.mlr.press/v206/hegselmann23a.html

15. Peng C, Yang X, Chen A, Smith KE, PourNejatian N, Costa AB, et al. A study of generative large language model for medical research and healthcare. NPJ Digit Med. 2023 Nov 16;6:210.

16. Dettmers T, Pagnoni A, Holtzman A, Zettlemoyer L. QLoRA: Efficient Finetuning of Quantized LLMs [Internet]. arXiv; 2023 [cited 2023 Sep 5]. Available from: http://arxiv.org/abs/2305.14314

17. He P, Gao J, Chen W. DeBERTaV3: Improving DeBERTa using ELECTRA-Style Pre-Training with Gradient-Disentangled Embedding Sharing [Internet]. arXiv; 2023 [cited 2024 Mar 18]. Available from: http://arxiv.org/abs/2111.09543

18. Liu NF, Lin K, Hewitt J, Paranjape A, Bevilacqua M, Petroni F, et al. Lost in the Middle: How Language Models Use Long Contexts [Internet]. arXiv; 2023 [cited 2024 Mar 18]. Available from: http://arxiv.org/abs/2307.03172

19. Singhal K, Tu T, Gottweis J, Sayres R, Wulczyn E, Hou L, et al. Towards Expert-Level Medical Question Answering with Large Language Models [Internet]. arXiv; 2023 [cited 2023 Aug 22]. Available from: http://arxiv.org/abs/2305.09617

20. Zhang H, Chen J, Jiang F, Yu F, Chen Z, Li J, et al. HuatuoGPT, towards Taming Language Model to Be a Doctor [Internet]. arXiv; 2023 [cited 2024 Mar 18]. Available from: http://arxiv.org/abs/2305.15075

21. Li F, Jin Y, Liu W, Rawat BPS, Cai P, Yu H. Fine-Tuning Bidirectional Encoder Representations From Transformers (BERT)–Based Models on Large-Scale Electronic Health Record Notes: An Empirical Study. JMIR Med Inform. 2019 Sep 12;7(3):e14830.

22. Sushil M, Ludwig D, Butte AJ, Rudrapatna VA. Developing a general-purpose clinical language inference model from a large corpus of clinical notes [Internet]. arXiv; 2022 [cited 2024 Mar 18]. Available from: http://arxiv.org/abs/2210.06566

23. Yang X, PourNejatian N, Shin HC, Smith KE, Parisien C, Compas C, et al. GatorTron: A Large Language Model for Clinical Natural Language Processing [Internet]. medRxiv; 2022 [cited 2024 Mar 18]. p. 2022.02.27.22271257. Available from: https://www.medrxiv.org/content/10.1101/2022.02.27.22271257v2

24. MIMIC-III, a freely accessible critical care database | Scientific Data [Internet]. [cited 2024 Mar 18]. Available from: https://www.nature.com/articles/sdata201635

25. Wornow M, Xu Y, Thapa R, Patel B, Steinberg E, Fleming S, et al. The shaky foundations of large language models and foundation models for electronic health records. Npj Digit Med. 2023 Jul 29;6(1):1–10.

26. Tworkowski S, Staniszewski K, Pacek M, Wu Y, Michalewski H, Miłoś P. Focused Transformer: Contrastive Training for Context Scaling [Internet]. arXiv; 2023 [cited 2024 Mar 18]. Available from: http://arxiv.org/abs/2307.03170

